# From Injury to Independence: Longitudinal Locomotor Recovery Patterns Following Traumatic Brain Injury - a TBI Model Systems Study

**DOI:** 10.64898/2026.07.07.26357255

**Authors:** Matthew J. Beth, Jennifer Marwitz, William Murrah, Nojan Valadi, Niyati Baweja, Harsimran S Baweja

## Abstract

**Background/Objectives:** Traumatic Brain Injuries (TBIs) affect more than 50 million individuals worldwide each year. Approximately 90% of individuals survive and experience persistent motor, cognitive, and emotional deficits, substantially contributing to a reduced quality of life and a global economic burden. TBI mechanisms are a foundational determinant of long-term recovery. The objective of this study was to examine long-term trends in functional locomotion ability over extended follow-up durations (>10 years) across distinct TBI mechanisms. The researchers hypothesized that TBIs caused by falls or violent mechanisms would be associated with poorer functional locomotor abilities and, subsequently, lower item scores than those sustained through automotive or recreational activities.

**Methods:** Data were obtained from the Traumatic Brain Injury Model Systems (TBIMS) database at Craig Hospital in Englewood, Colorado, the world’s largest longitudinal TBI data repository. Functional locomotion was assessed using the Functional Independence Measure (FIM) Locomotion item as the primary outcome measure. To enhance measurement precision and ensure interval-level scaling, raw FIM scores were converted into logit-based estimates of latent functional ability using Rasch modeling. Longitudinal changes of these Rasch-transformed scores were analyzed using linear mixed-effects regression, accounting for individual-level variability and unbalanced follow-up data.

**Results:** The findings demonstrated a clinically meaningful decline in functional ability among individuals with TBIs from violent mechanisms, particularly assault-related injuries and gunshot wounds, which were associated with chronic medical complications and limited functional independence. Conversely, TBIs from bicycling, unclassified vehicular incidents, and winter sports showed significant positive estimates, possibly reflecting higher premorbid physical fitness. Motor vehicle, motorcycle, pedestrian, and fall-related TBIs demonstrated steep early gains, followed by a period of recovery stabilization and plateau. In contrast, violence-related mechanisms were characterized by consistently low median scores, with minimal long-term improvement. Falls, gymnastics, track & field, and water sports did not exhibit meaningful changes in the context of the primary hypothesis.

**Conclusions:** TBI mechanisms play a vital role in shaping long-term functional locomotion outcomes, with violence-related TBIs associated with poorer long-term functional independence. The results have clinically important implications, supporting earlier identification of high-risk populations and the development of targeted rehabilitation strategies during periods of heightened neuroplasticity. Rasch analysis integrated with linear mixed-effects modeling yields a robust analytic framework that uncovers subtle but meaningful differences in recovery trajectories across TBI mechanisms.

## INTRODUCTION

Globally, more than 50 million Traumatic Brain Injuries (TBIs) are sustained annually (1). Only approximately 10% of TBIs sustained are fatal, meaning that 90% of individuals who suffer TBIs, or about 45 million people each year, survive their injuries and live with residual disabilities (2). As of 2021, TBIs accounted for approximately 5.48 million years lived with disability worldwide (3). The chronic disabilities that survivors of TBI experience often include persistent motor, cognitive, and/or emotional impairments.

Additionally, TBIs incur a substantial economic burden. Individuals with moderate to severe TBIs often require prolonged hospital stays and intensive rehabilitation services. A review of TBI rehabilitation explains that important and expensive interventions, including extended hospitalizations and inpatient rehabilitation, are often necessary and account for significant expenditures (4). Long-term healthcare needs significantly impact global healthcare expenditures. Reports estimate that TBIs cost over 400 billion USD globally each year (5). One severe TBI alone incurs a cost between 2,130 and 401,808 USD for in-hospital expenses only, and expenditures steepen further from long-term outpatient rehabilitation and adjusting to life with newly acquired disabilities from their brain injury (6).

Many adjustments are accommodations for impaired independence in functional activities resulting from their injuries (7). In addition to the exorbitant medical costs, poorer functional independence reduces long-term quality of life (8) and places a significant burden on families and society (9,10). Therefore, it is imperative to investigate reduced functional independence.

### Falls

A strong association exists between the mechanism of TBI and long-term recovery outcomes, showing that individuals with TBIs due to falls consistently have poorer long-term outcomes compared to their peers injured by other mechanisms (11). Meanwhile, TBIs from road traffic accidents and sports-related activities are associated with poor outcomes, but to a lesser extent than those from TBIs from falls (11). Researchers later added to this, finding that traffic-related mechanisms of TBI often cause the most severe injuries, yet are associated with more favorable outcomes (12). In contrast, fall-related mechanisms of injury often cause less severe injuries, yet are associated with worse outcomes (12).

### Injury Severity

Latent class mixed models identified low, medium, and high trajectories for the cognitive and motor domains of the Functional Independence Measure (FIM). Multinomial logit models created predictive odds ratios for group membership in the latent classes. The analysis showed that open head injuries and extended durations between injury onset and rehabilitation admission were both indicative of a worse motor recovery trajectory (13).

A study following TBI survivors for 24 years post-injury revealed that the population experienced significant functional limitations even after extended follow-up periods. The study found that injury severity and TBI mechanism were positively associated with limitations (14). In another study, 47.5% of a sample of 504 individuals reported experiencing balance difficulties 8 years post-TBI. Additionally, 31% of the sample expressed having problematic motor deficits impairing their ability to move their body or extremities after eight years post-injury (15). The analysis revealed an inverse correlation between injury severity and long-term functional outcomes, suggesting that more severe injuries are associated with worse long-term outcomes. The study further reported that only 48.7% of participants (245 participants) returned to work and were employed full-time. Further, 38% of participants (191 participants) reported that their salary had been reduced post-TBI, primarily due to performance limitations resulting from TBI-related impairments (15).

A latent profile analysis identified four latent classes, each with linearly increasing with greater functional impairment. The analysis determined that the LPA class four showed the greatest impairment. Characteristics typifying LPA class four were the most severe TBIs, including significant functional impairment, cognitive decline, and critically impaired consciousness, leading to poorer outcomes and more extended hospital stays (16). Additionally, a meta-analysis investigated the impact of injury severity on outcomes over the first year post-TBI reviewed studies with an average effect size of *r* = .26 and a 95% confidence interval of .20 to .31, indicating a moderate association between TBI severity and outcomes at 1 year post-TBI. The researchers examined heterogeneity across the included studies. Thus, Cochran’s *Q* and *I²* statistics were key metrics in their meta-analysis. Their meta-analysis revealed *Q*(27) = 381.75 and *p* < .00001, and an *I²* value of 93.1%. This indicates a high degree of variability in effect sizes across the studies included in the meta-analysis. Ultimately, the study indicated that injury severity significantly predicts outcomes one year after the injury (17).

### Falls, Violence, & Injury Severity

A retrospective study found that TBI severity was significantly associated with mechanism of injury: falls from height and gunshot injuries were among the mechanisms linked to severe TBIs (18). Gunshot wounds and open-head injuries accounted for the highest percentage of severe TBIs in a sample of 401 participants (55.7%) (18). Many individuals with moderate to severe TBI had gross neuromotor impairments, including persistent difficulties with walking, balance, postural control, spastic paralysis, impaired limb control and motor coordination, and reduced fine motor control years post-injury (19). Individuals with open head injuries and injuries from acts of violence carry a higher risk of severe damage, associated hemorrhages, fractures, cerebrospinal fluid leaks, and other complications (20), all of which are indicative of more severe injuries.

With these findings, it is evident that more severe TBIs were likely a result of a fall or an act of violence. Thereby, the injury severity findings can reasonably be applied to TBIs caused by falls from height, gunshot wounds, blast injuries, and assaults.

### Traumatic Brain Injury Model Systems

The data used in this analysis were obtained from the Traumatic Brain Injury Model Systems (TBIMS) database at Craig Hospital in Englewood, Colorado. The TBIMS was founded in 1987, and its data are used for research, quality improvement initiatives, and to inform clinical practice in TBI rehabilitation (21). Researchers have identified the TBIMS as the world’s largest longitudinal Traumatic Brain Injury database (22). The database contains data on more than 20,000 individuals with TBI collected over nearly 40 years (23).

The TBIMS collects a comprehensive dataset, including data regarding the Disability Rating Scale (24,25), the FIM, substance use history, arrests, and psychiatric problems within the previous year, rehospitalizations, and social and global outcomes from the Glasgow Outcome Scale–Extended (26,27), the Supervision Rating Scale (28), the PART- O participation instrument (29), and the Satisfaction with Life Scale (30). Data are collected at rehabilitation admission and discharge, and follow-up data are collected via telephone interviews at the first, second, fifth, and tenth injury anniversaries, and every 5 years thereafter (31).

The TBIMS reports the mechanism of injury with the “Cause” variable. The variable has 19 levels (representing motor vehicle, motorcycle, bicycle, all-terrain vehicle, other vehicular unclassified, gunshot wound, assault with blunt instrument, other violence, water sports, field/track sports, gymnastic activities, winter sports, air sports, other sports, fall, hit by falling/flying object, pedestrian, and other unclassified).

### Functional Independence Measure

The data in this study were collected using the Functional Independence Measure (FIM) (32,33). The FIM is an 18-item instrument that quantifies the burden of care relating to motor (13 items) and cognitive (5 items) functioning. Items are rated on a 7-point scale, ranging from total assistance (1) to complete independence (7) (34). The FIM evaluates problem-solving, memory, expression, comprehension, social engagement, eating, bathing, grooming, upper-body dressing, lower-body dressing, toileting, bowel and bladder management, transfers (bed, chair, wheelchair, toilet, and shower/bathtub), locomotion, and stair climbing (34). At admission and during in-person rehabilitation, the FIM is administered by professional clinicians who observe an individual’s functioning across the various scenarios and tasks assessed. Follow-up data are primarily collected via telephone interviews (31).

Use of the FIM in head injury rehabilitation has been studied for decades. In 1998, the use of the FIM in neurotrauma rehabilitation settings yielded an inter-rater reliability coefficient of *r* = .95, with the motor subscale showing the highest reliability (35). Additionally, in 2016, the FIM indicated that the instrument exhibited high internal consistency, yielding an alpha value of *α* = .98 (34).

### Rasch Analysis

FIM data are susceptible to ceiling effects, making it imperative to employ Item Response Theory (IRT) methods prior to analysis. A Rasch analysis satisfies the IRT necessity, transforming the raw ordinal data into interval-level logit scores via the partial credit model. The conversion is paramount because linear models assume equally spaced data, and violations of this assumption can bias estimates and invalidate standard errors. Meanwhile, the difference between the raw scores of 3 for “moderate assistance” and 4 for “minimal assistance” is not necessarily equal to the difference between scores of 5 for “supervision” and 6 for “modified independence.” Rasch analyses convert the raw 1–7 item scores into a continuous, evenly spaced Rasch logits. This means that the distance between scores of “3” and “4” is now equal to the distance between scores of “5” and “6”. Although the original data represent ordinal FIM Locomotion scores, the transformation evaluates the underlying latent locomotion ability. In addition to creating equal intervals, the transformation improves data linearity and residual normality, enhancing the power and accuracy of fixed- and random-effects estimates, and legitimizing group comparisons.

Despite growing evidence that TBI outcomes vary by mechanism of injury, previous studies have primarily focused on how TBI mechanisms relate to short-term functional outcomes (typically 1-8 years) and have often used raw FIM scores in analyses, which may obscure meaningful differences across groups. As a result, the impact of the mechanism of injury on short-term functional outcomes is relatively well understood, but this leaves a critical gap in understanding whether mechanisms of injury are associated with distinct recovery trajectories when FIM locomotion scores are measured more precisely using Rasch-transformed scores. To address this gap, the present study uses linear mixed-effects regression to evaluate longitudinal patterns in FIM locomotion scores across TBI mechanisms. Specifically, we ask whether Rasch-transformed FIM locomotion scores over extended periods (+10 years) will vary by TBI mechanism. We hypothesize that individuals with TBIs caused by falls or violence-related mechanisms of injury will show lower FIM locomotion scores (representing locomotor functioning) compared to individuals with TBIs from automotive or recreational mechanisms of injury.

## METHODS

### Participants

Secondary data from the Traumatic Brain Injury Model Systems (TBIMS) national database were used in this study. The TBIMS dataset contains de-identified data for 30,768 individuals. However, the study examined the impact of the mechanism of injury on very long-term locomotor functional independence, and the analysis excluded participants with fewer than 10 years of recorded data. The exclusion reduced the total sample size to 6,226.

### Statistical Analysis

The study conducted a linear mixed-effects regression to examine predictors of locomotor functional independence. The model included two fixed effects: the first fixed effect was the TBI mechanism (Cause), a nominal predictor with 19 levels (motor vehicles, motorcycles, bicycles, all-terrain vehicles/cycles, unclassified vehicular accidents, gunshot wounds, assaults with blunt instrument, other violence, water sports, track sports, gymnastic activities, winter sports, air sports, other sports, falls, hit by falling/flying objects, pedestrian-related injuries, and other unclassified injuries). The second fixed effect was an interaction term (AGENoPHIFxDAYStoFUF), combining the variables for age (AGENoPHIF) and days to follow-up (DAYStoFUF). The purpose of this term in the model was to investigate whether longitudinal recovery trajectories varied by a participant’s age.

The model’s random effects, accounting for hierarchical clustering, included the participant identifier (Mod1Id) and the geographic location of residence just before injury (ZipInj). Variance components for the random factors were estimated. The “FIMLocoF” variable served as the outcome variable in the model, rather than “FIMLoco,” because “FIMLocoF” represents the data collected across the longitudinal follow-ups.

The statistical significance of the fixed effect was assessed using t-tests on parameter estimates, and an F-test assessed the overall significance of the “Cause” variable. The alpha level, or threshold for statistical significance, was set at 0.05 (36). All statistical analyses were performed using R (v4.5.1) (37). The Rasch analysis was conducted using the ’TAM’ R package (38), and the ’mosaic’ R package (39) was used to calculate descriptive statistics for the FIMLocoF variable, both as raw scores and as Rasch-scaled scores. The values are listed in Table 1. After the data were converted to logit scores, a linear mixed-effects regression was used to evaluate the relationships in the study. The study used the’lme4’ R package (40) to fit mixed models and the ’ggplot2’ R package (41) to create figures.

**Table 1.**
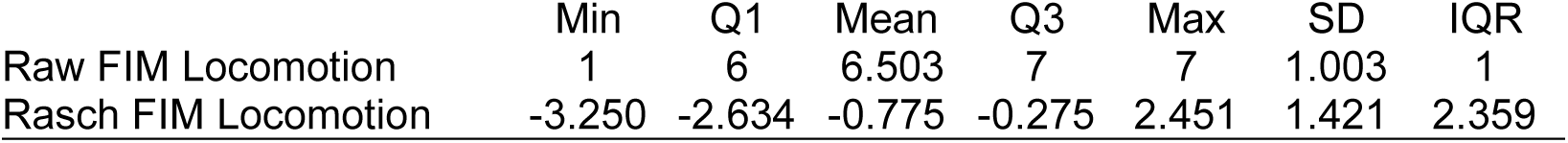
Descriptive statistics for the raw FIM Locomotion scores and the logit FIM Locomotion scores after Rasch analysis.

**Table 2.**
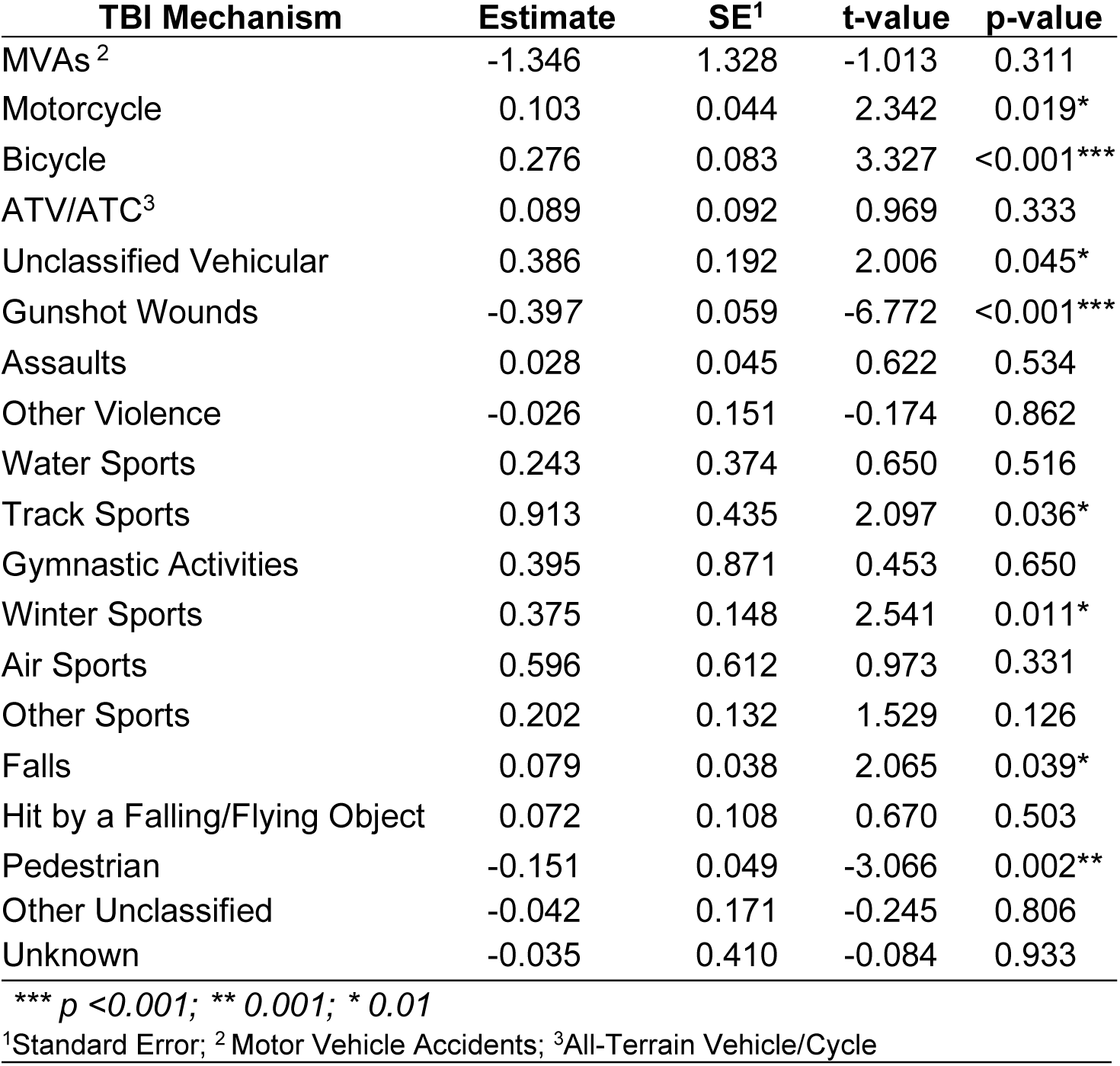
Summary and coefficient estimates from the fixed effect Cause variable in the linear mixed-effects regression with the Rasch-FIM Locomotion scores as the outcome variable.

## RESULTS

The study fit a linear mixed-effects model to examine the associations between Rasch-FIMLocoF scores (Rasch transformed follow-up FIM Locomotion scores), with the TBI mechanism (Cause) and an interaction term between age (AGENoPHIF) and days until follow-up (DAYStoFUF) as the fixed effects, while considering random effects, from participant identifier (Mod1Id) and pre-injury geographic location of residence (ZipInj).

The linear mixed-effects model identified significant fixed effects for the TBI mechanism, age at the time of injury, follow-up time, and the interaction between age and follow-up time (see Table 3). The findings should instill confidence in the robustness of the results, reassuring researchers and clinicians of the reliability of the observed variations in functional locomotion outcomes across TBI mechanism groups over time.

**Table 3.** Analysis of Variance (ANOVA) table of the components of the model’s fixed effects.

|  | Sum Sq | Mean Sq | NumDF | DenDF | F Value | Pr(>F) |
| --- | --- | --- | --- | --- | --- | --- |
| Cause | 149.92 | 8.329 | 18 | 5439.1 | 5.797 | 5.038e-14*** |
| AGENoPHIF | 186.12 | 2.482 | 75 | 25068.6 | 1.727 | 9.807e-05 *** |
| DAYStoFUF | 19.28 | 19.283 | 1 | 25160.8 | 13.420 | .0002495 *** |
| AGENoPHIFxDAYStoFUF | 377.54 | 5.172 | 73 | 25860.1 | 3.599 | < 2.2e-16 *** |

\*\*\* $p < 0.001$ ; \*\* $0.001$ ; \* $0.01$

Several TBI mechanisms were significantly associated with very long-term functional locomotion, thereby informing targeted rehabilitation strategies and clinical decision-making.

At the omnibus level, the age-at-injury by follow-up time interaction was statistically significant, supporting the inference that recovery trajectories varied by age across the follow-up data collections. However, when interaction coefficients were examined individually, effect sizes were small, and statistical significance across specific age categories was borderline. Most age-by-time interaction estimates ranged from −0.001 to −0.002, with *p*-values exceeding .05, suggesting the interaction reflected a broad cumulative pattern rather than sizable differences between age groups. For example, among older age groups, interaction estimates for AGENoPHIF67 × DAYStoFUF (Estimate = −0.00149, SE = 0.00128, *t* = −1.17, *p* = .242), AGENoPHIF77 × DAYStoFUF (Estimate = −0.00153, SE = 0.00128, *t* = −1.20, *p* = .232), and AGENoPHIF81 × DAYStoFUF (Estimate = −0.00153, SE = 0.00128, *t* = −1.20, *p* = .231) each of which did not meet the required level of statistical significance.

Model fit statistics indicated adequate performance for the functional locomotion outcome. Random-effects estimates revealed notable between-participant variability, reinforcing the rationale for participant-level random intercepts. Variance attributable to pre-injury residential zip code indicated modest geographic clustering, whereas the residual variance accounted for a significantly larger proportion of the variability, suggesting that most unexplained variation was concentrated at the inter-individual level, rather than at the regional level.

## DISCUSSION

The objective of the study was to examine the differential impact of TBI mechanisms on very long-term (+10 years) FIM locomotion scores. The TBI mechanism strongly predicts long-term locomotor functional independence, with an F value of 5.797 and *p* <.0001 (see Table 3). The study found statistically significant negative estimates for individuals with TBIs caused by gunshot wounds and pedestrian-related accidents. Significant negative estimates are presented in Figure 1. Injuries from gunshot wounds are consistently associated with the lowest functional scores over extended periods. Like other mechanisms of injury, TBIs caused by gunshot wounds are characterized by a myriad of initial health complications, including hypoxia, ischemia, elevated intracranial pressure, and cranial nerve injuries. However, gunshot wounds also pose risks to individuals with TBI long after the initial injury. When symptoms are categorized into early, intermediate, and late phases, these individuals may also experience complications such as seizures, refractory cerebral edema, acute hydrocephalus, vasospasm, cerebrospinal fluid leaks, deep venous thrombosis, and pseudoaneurysms during the intermediate phase (42). During the late phase, individuals with TBIs from gunshot wounds may also be challenged with conditions like infections, hydrocephalus, cerebrospinal fluid fistula, venous sinus occlusions, trephination syndrome, scalp necrosis, arteriovenous fistulas, lead/copper toxicity from bullet fragments, temporalis atrophy, and hygroma (42). The chronic health complications the population faces are a primary contributor to the lower scores and functional independence.

**Figure 1.**
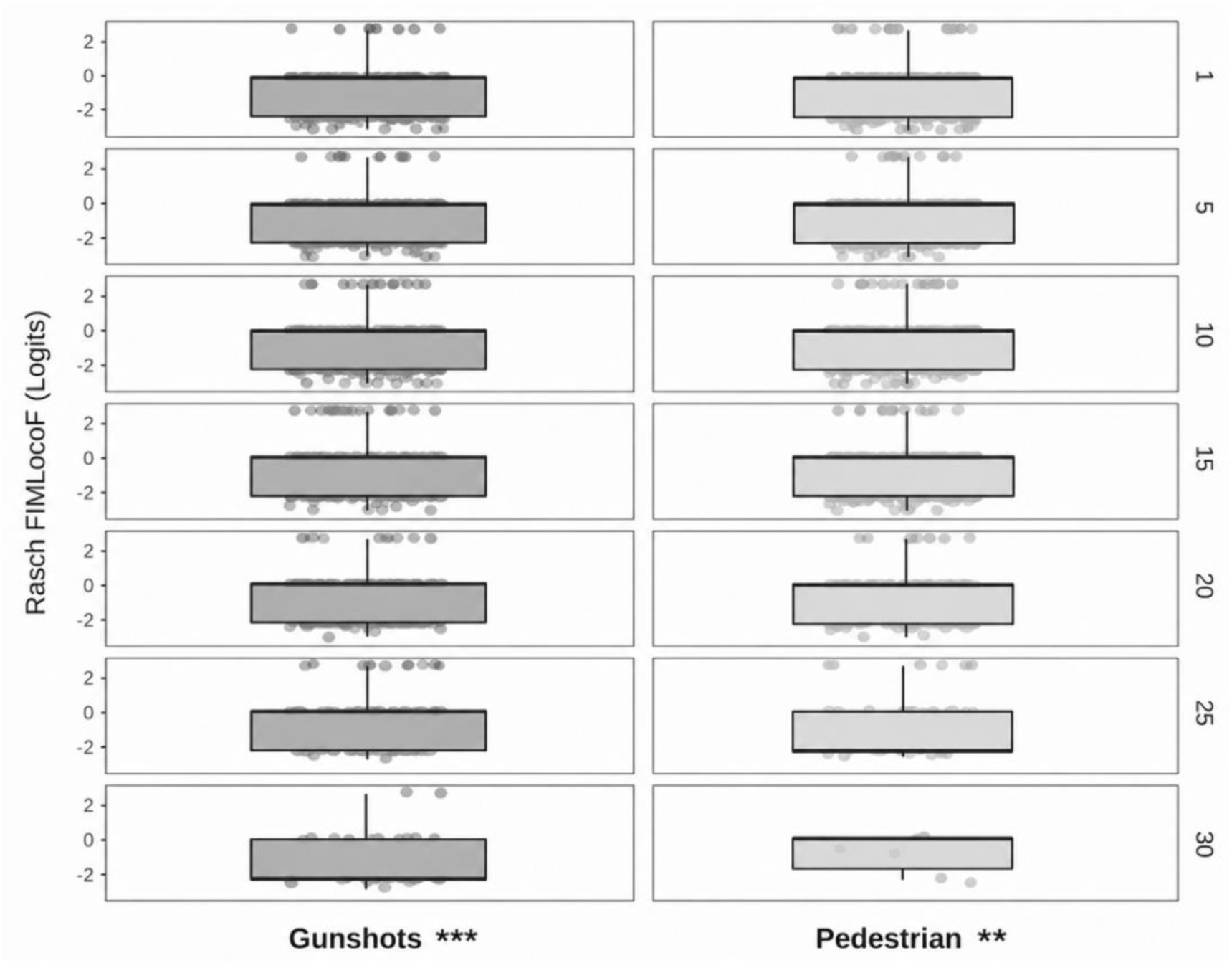
Longitudinal distributions of Rasch-transformed FIM Locomotion scores by TBI mechanism. Each column represents individuals with different TBI mechanisms (gunshots and pedestrian-related injuries), with significant negative estimates. Each row corresponds to the Rasch-transformed FIM Locomotion scores at subsequent follow-up assessments. Box plots depict the median and interquartile range, with individual points representing participant observations. Across follow-ups, pedestrian injuries demonstrate consistently higher locomotor functioning, whereas gunshot and other violence mechanisms consistently show the poorest scores. Motor vehicle and assault-related TBIs display moderate outcomes, with substantial variability, indicating enduring differences in locomotor recovery trajectories by TBI mechanism. *p* <0.001 *** ; 0.001 ** ; 0.01 *

Conversely, statistically significant positive estimates were found for individuals with TBIs caused by bicycles, unclassified vehicular, falls, and both track and winter sports. Individuals with injuries from bicycles, track, and winter sports likely have a more athletic background and may be more physically fit. Current research using human models showing that exercise before TBI enhances recovery outcomes is limited (43). However, animal models provide strong evidence supporting the previous finding that fitness supplements improve TBI recovery. Research in mice conditioned prior to TBI found that voluntary exercise before injury significantly enhanced post-injury outcomes (44). Additionally, the exercised mice brains showed elevated expression of neuroprotective growth factors, including VEGF-A and EPO (44). The adaptations would enable a more physically fit population to more successfully regain their functional abilities over time.

TBI mechanisms, including motorcycle, all-terrain vehicle (ATV/ATC), water sports, field/track sports, gymnastic activities, air sports (e.g., parachuting), other sports, falls, being hit by a falling/flying object, and other unclassified causes, did not affect individuals’ ability to recover their functional locomotor abilities. The variables Mod1Id and AGENoPHIF were statistically significant random effects. However, the residual variance accounted for most of the variance, suggesting that most of the unexplained variability in locomotion function remains within the individual participants after accounting for fixed and random effects. The study’s findings confirmed a portion of the hypothesis and refuted another part. Average Rash-transformed FIM-LocoF scores over 30 years for TBI mechanisms related to the study hypothesis are shown in Figure 2. The TBI mechanisms depicted include motor vehicles, motorcycles, ATVs & ATCs, unclassified vehicular, gunshots, assaults, other violence, falls, track and field, gymnastics, water, winter, air, and other sports. The study observed a meaningful decline in the functional ability of individuals with TBIs from gunshots and meaningful functional locomotor improvements in individuals with TBIs from motorcycles, unclassified vehicular accidents, falls, track and field, and winter sports. However, the study did not observe a meaningful change in the functional ability of individuals with TBIs from motor vehicles, ATVs & ATCs, assaults, other violence, gymnastics, water, air, and other sports.

**Figure 2.**
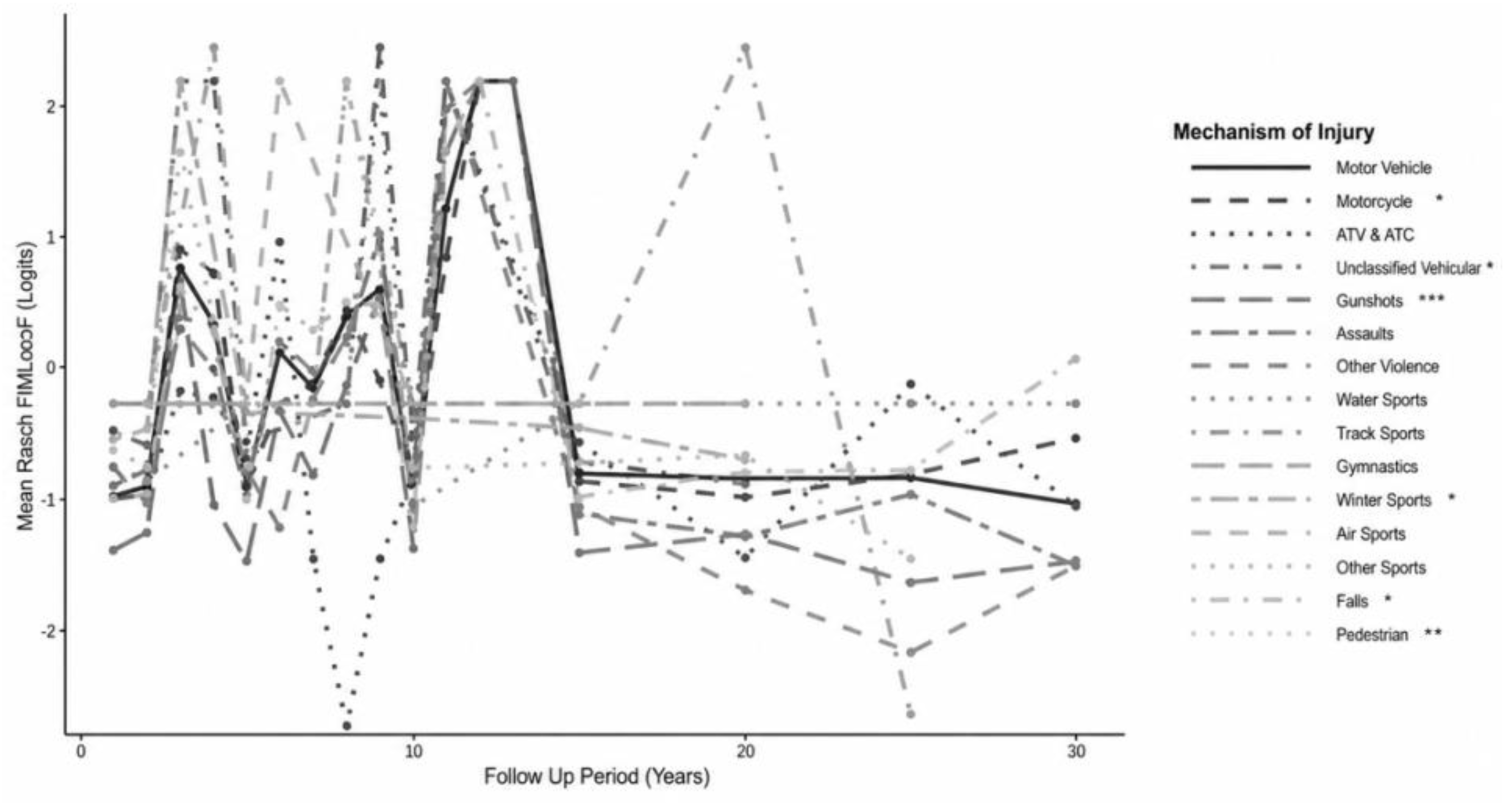
Trajectories of Mean Rasch-FIM Locomotion Scores by Mechanism of TBI Across Follow-Up Periods. Mean Rasch-transformed Functional Independence Measure (FIM) Locomotion scores (logits) across 30 years for different TBI mechanisms. Each line represents an injury mechanism, differentiated by line type. The Y-axis indicates the average Rasch-transformed FIMLocoF scores (or the latent locomotion ability) per TBI mechanism, with logit values ranging from −3.25 to 2.45. Higher logit values indicate better locomotor functioning. Rasch-FIM Locomotion scores show significant variability during earlier years, whereas scores converge toward lower, negative logit values during later years, suggesting long-term locomotor impairments across TBI mechanisms. *** *p* <0.001; ** 0.001; * 0.01

### Rasch Interpretation

A Rasch analysis transformed the raw, ordinal Functional Independence Measure (FIM) scores into interval-level, logit-based estimates of functional ability. The logits measure underlying functional capacity, enabling score changes to be interpreted as equal units of functional change across an instrument’s measurements. As such, the coefficients from the linear mixed-effects models reflect meaningful differences in latent functional ability rather than in raw, ordinal scores.

The transformation strengthens the validity of longitudinal modeling and supports a more accurate interpretation of recovery trajectories across injury mechanisms. As such, a change of 0.5–0.7 logits is considered moderate in Rasch applications (45). The raw FIM Locomotion item was scored initially on a 1–7 ordinal scale. After Rasch scaling, logit values in this sample ranged from −3.25 to 2.45 logits (*M* = −0.78 logits), with an interquartile range of 2.36 logits (*Q1* = −2.63 logits, *Q3* = −0.28 logits) (see Table 1).

### AGENoPHIF×DAYStoFUF Interaction

The study findings revealed a meaningful interaction between age at injury (AGENoPHIF) and follow-up time (DAYStoFUF) in predicting locomotor functional independence. The AGENoPHIF × DAYStoFUF interaction reached statistical significance, *F*(73, 25,860.1) = 3.60, *p* < .001 (see Table 3), confirming that the relationship between time post-TBI and locomotor recovery varies across age groups. While most individual interaction coefficients were modest and did not reach statistical significance, the overall pattern underscores the importance of accounting for age in long-term recovery trajectories.

Negative interaction coefficients were predominant at older ages, indicating that longer follow-up periods were associated with smaller functional gains and greater declines in locomotion among older individuals compared to younger ones. This underscores the importance of considering age-related factors, such as diminished neuroplasticity, comorbidity burden, and reduced physiologic reserve, which are well-established in TBI rehabilitation literature. Consequently, older adults may experience slower locomotor recovery and face challenges maintaining long-term mobility-related independence post-TBI.

The interaction suggests that age should be viewed as a dynamic factor influencing post-TBI recovery over time, rather than a static variable. Clinically, these results support the need for long-term, individualized rehabilitation plans and ongoing monitoring, especially for older patients, as their recovery trajectories may diverge significantly from those of younger individuals. Tailoring mobility and locomotor interventions over extended periods can optimize outcomes based on age-related recovery patterns.

However, the interaction yielded coefficients of modest magnitude, pointing to a gradual rather than a sharp effect. Multiple variables influence functional locomotion outcomes beyond age, including the TBI mechanism, adequate healthcare and rehabilitation exposure, and psychosocial and environmental factors. Regardless of its magnitude, the significant interaction supports incorporating time-dependent age-related effects into future prognostic frameworks for predicting long-term functional mobility outcomes post-TBI.

### Recovery Trajectories

Figure 2 depicts the mean Rasch-FIM Locomotion scores for different TBI mechanisms related to the study hypothesis over 30 years. Latent Locomotion abilities vary widely across mechanisms during the early follow-up years, with some TBI mechanisms showing scores well above 0 logits and others below, but the differences diminish over time. After 10–15 years post-TBI, trajectories flatten and converge towards negative values, with most TBI mechanisms between −0.5 and −1.5 logits, indicating a long-term plateau or decline rather than sustained improvement. Mechanism-related differences are evident early, suggesting that injury mechanism strongly influences early functional recovery, whereas long-term functional locomotion becomes more similar across TBI mechanisms. Vehicular injuries are associated with higher locomotion scores during the early follow-up years, followed by a subsequent decline, indicating early functional gains that later deteriorate. Violence-related mechanisms (i.e., gunshots, assaults, and other violence) have low initial scores, which gradually decline further over time, reflecting consistently poor long-term functional outcomes. Conversely, sports-related TBIs show significant variability, with peaks in the early follow-up years that converge toward negative logits later. Falls are relatively stable, with lower-magnitude peaks and valleys, mostly hovering around slightly negative logits over time. Interpreted in Rasch terms, where differences between 0.5–1.0 logits represent clinically meaningful variation in functional ability (45), the widespread shift into negative logits during later follow-up years indicates enduring locomotor impairments decades after TBI across mechanisms.

Figure 3 presents a matrix of boxplots of Rasch-FIMLocoF scores for individuals, stratified by TBI mechanism, across various follow-up periods. During earlier follow-up periods, TBIs from motor vehicles, motorcycles, pedestrian-related injuries, and falls show steeper improvements in median FIM Locomotion scores that subsequently plateau, indicating that these individuals regain considerably more ambulatory ability earlier in their recovery. This aligns with the known recovery declines associated with increases in time post-injury and normative aging.

**Figure 3.**
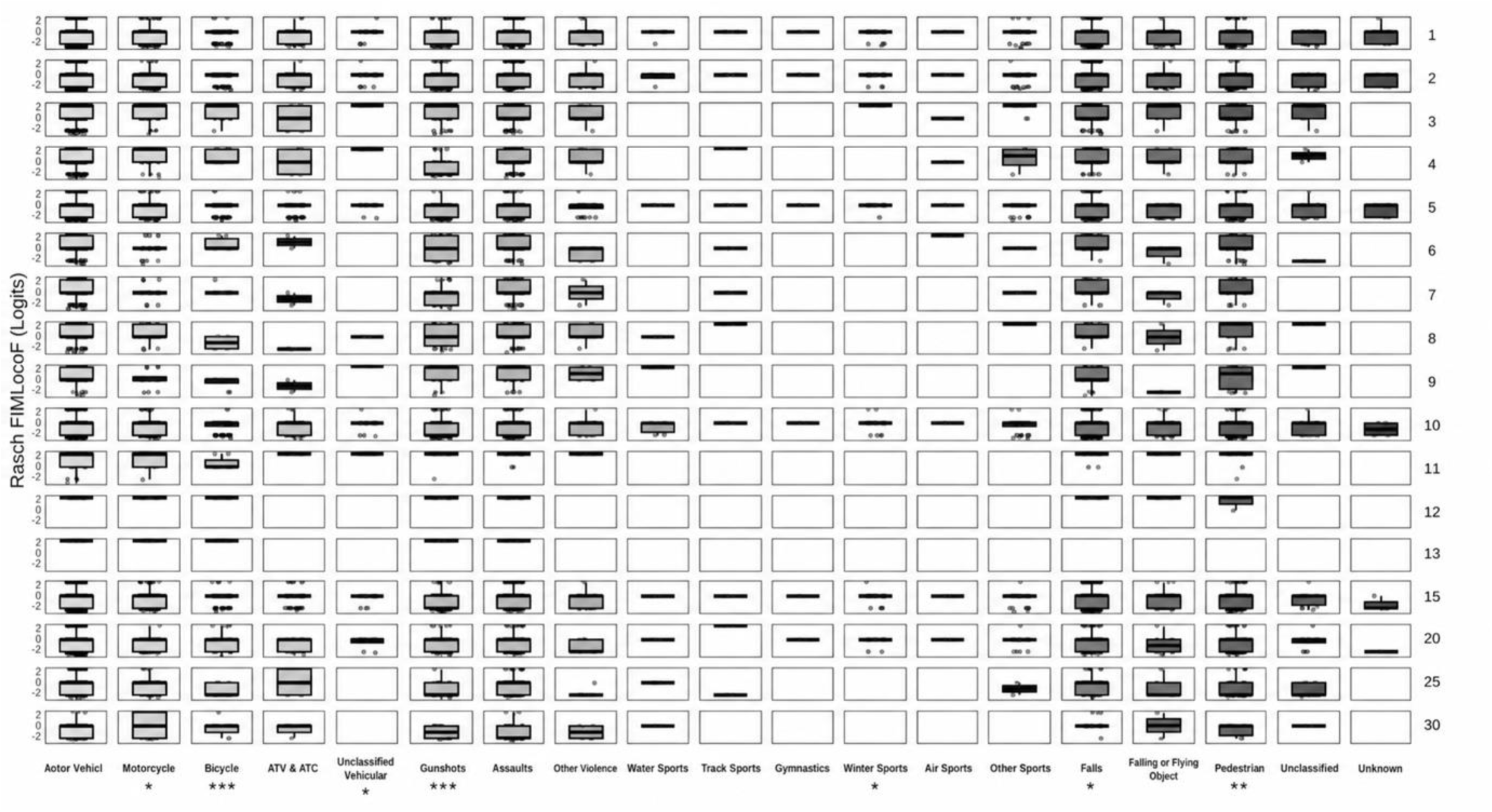
Longitudinal Trajectories of Rasch-Transformed FIM Locomotion Scores by Mechanism of Injury; Box plots depict the distribution of Rasch-transformed Functional Independence Measure (FIM) Locomotion scores across follow-up time points for individuals with TBI, stratified by TBI mechanism (columns). Rows represent the measure at each follow-up period. Recovery trajectories differ by injury mechanism, with TBIs from gunshots showing relatively lower functional independence and minimal improvement over time. *** *p* <0.001; ** 0.001; * 0.01

Individuals with TBIs caused by gunshots, assaults, and other violence demonstrate relatively low early median FIM Locomotion scores that show little improvement over time. The pattern suggests that these TBI mechanisms often result in severe, long-lasting locomotor deficits with limited functional improvement over time. Lastly, TBIs resulting from bicycles, water, track, gymnastics, winter, and air sports show high median FIM Locomotion scores at early follow-up periods. The trajectories for these injury mechanisms are nearly flat, as there is little room for improvement.

### Significance and Implications

The study findings have significant clinical and practical implications. With knowledge of how TBI mechanisms affect functional outcomes, clinicians can quickly discern, even at the site of injury, which tasks may be disproportionately challenging for a given individual and require additional therapeutic attention. Rapidly learning this information enables researchers and practitioners to design and implement targeted TBI rehabilitation interventions more quickly. Quickly implementing therapeutic interventions allows individuals with TBI to participate in the therapies and begin their rehabilitation journey while still benefiting from their critical period, the time most neurologically advantageous for recovery from TBI, and affords individuals with TBI the best opportunity to recover from their newly acquired impairments.

The critical period is short-lived, as the optimal effects occur within the first 6 months post-injury. After the initial 6 months, the beneficial neuroplasticity window gradually closes between 6 and 24 months post-injury. A cascade of cellular and molecular processes characterizes this period, including increased axonal sprouting, dendritic remodeling, and synaptogenesis, as well as upregulation of NMDA receptors and downregulation of GABA_A_ receptors (46,47).

Findings from this study reveal tasks that may be disproportionately difficult for TBI survivors of a given mechanism of injury. The knowledge gained from these results will inform future research to optimize TBI rehabilitation outcomes and improve the lives of individuals with TBI.

Additionally, the findings help inform long-term care needs and resource allocation. The study discusses functional recovery trajectories, as depicted in Figure 3, providing individuals with TBI prognostic insights into their recovery journey.

Knowing this information will help determine which future activities may be handled independently and which may require additional assistance. The findings may also inform resource allocation. A snowy, mountainous region where skiing and snowboarding are common activities should be particularly mindful to ensure adequate staffing of neurosurgeons, orthopedic surgeons, and other critical health care professionals. These professionals may need specialized equipment, advanced imaging technology, and ventilators in the area. Furthermore, therapeutic services in the physical, occupational, and speech-language pathology domains must be available in the region. Winter sports show high median FIM Locomotion scores with minimal further improvement needed, suggesting that the population maintains high-level functioning throughout their recovery. As such, return-to-work and return-to-school practices may be more common in the specific region, and therefore, a higher prioritization of occupational therapy would be necessary. The region may have a greater demand for assistive devices such as wheelchairs, mobility aids, and Augmentative & Alternative Communication (AAC) devices than nearby areas, and additional funding and/or resources may be required to meet these needs.

### Previous Literature

The present findings differ in multiple ways from previous research reporting on differences in functional outcomes across TBI mechanisms. First, sample characteristics likely contribute to differences, as the current cohort was drawn from a national sample with heterogeneous TBI mechanisms and extended longitudinal follow-up data (over +10 years). In contrast, previous studies have often used smaller sample sizes over shorter follow-up periods (commonly between 1-8 years). Second, methodological heterogeneity in outcome measurement may account for results differences. Previous studies have analyzed and reported on raw, ordinal FIM scores, which are prone to strong ceiling effects. However, this study transformed the outcome variable using Rasch analysis, yielding FIM logits that provide an interval-level representation of functional ability. The transformation enables differences in outcome measures to be interpreted as equal units of functional change across the scale, revealing group differences that may be obscured when analyzing raw FIM scores. Lastly, heterogeneous statistical approaches may also contribute to any differences. This study uses linear mixed-effects modeling, which accommodates unbalanced longitudinal data and captures both fixed effects and individual-level variability in recovery trajectories. The varying methodological approaches may explain why this analysis detected significant, clinically meaningful differences in very long-term functional outcomes across injury mechanisms.

### Strengths & Limitations

Methodological strengths bolster the rigor and interpretability of the study findings. Firstly, using linear mixed-effects modeling enabled the analysis of longitudinal repeated functional outcome measures while accounting for between-group and between-individual variability in recovery trajectories. The analytical approach suits longitudinal TBI data well, as it accommodates unequal follow-up intervals and incomplete data without casewise deletion. Second, the Functional Independence Measure (FIM) Locomotion score data were transformed using Rasch analysis, yielding interval-level logit scores that better reflect underlying functional ability and were subsequently used in the mixed-effects models. This transformation mitigates ceiling and floor effects of ordinal raw FIM scores and improves comparability across items and time points, thereby supplementing the validity of longitudinal inferences. Another methodological strength was the use of multiple mechanisms of injury over extended follow-up periods, which enables a more nuanced assessment of long-term functional trajectories across heterogeneous mechanisms of TBI, yielding clinically meaningful results that extend far beyond short-term outcomes.

Despite the strengths, certain limitations warrant consideration. Although the overall sample size was sufficient for mixed-effects modeling, specific injury mechanisms (especially more obscure TBI mechanisms like gymnastics) were represented by relatively small subsamples, potentially limiting statistical power and generalizability for these mechanisms. Additionally, study participants with available long-term follow-up data may differ systematically from those lost to follow-up, potentially introducing selection bias. While Rasch transformations improve measurement functioning, they assume unidimensionality and local independence; if these assumptions are violated, score interpretation may be affected. From a modeling standpoint, the specified random-effects structure may not capture all sources of between-person heterogeneity, including confounding variables such as injury severity, substance abuse, and socioeconomic characteristics, which may contribute to observed differences across mechanisms. Finally, time since injury was modeled in discrete intervals rather than as a continuous process, potentially obscuring nonlinear recovery patterns.

### Future Research

The findings from this study provide a critical foundation for understanding the differential effects of TBI mechanisms on long-term functional locomotion outcomes. To further advance this understanding, future research should address specific methodological considerations. While the Traumatic Brain Injury Model Systems (TBIMS) is an extensive national database, uncommon and more obscure TBI mechanisms, such as TBIs caused by gymnastics and air sports, were represented by relatively small subsamples, thereby potentially limiting the generalizability and statistical power of these TBI mechanisms. Future studies may benefit from using different large samples to complement these findings and ensure robust, reliable analyses of less common TBI mechanisms. In addition, the study’s modeling of time since injury in discrete intervals, rather than as a continuous predictor, may not fully capture the complex, nonlinear nature of TBI rehabilitation. Exploring nonlinear growth curves, such as spline models, may yield a more nuanced and accurate depiction of functional changes over time.

Further, to provide a more holistic perspective on recovery from TBI, subsequent research should expand beyond motor functioning. The Functional Independence Measure (FIM) itself includes both motor and cognitive domains. Analyzing cognitive FIM scores would be a critical next step to understanding how injury mechanisms differentially affect cognitive functional independence, a significant component of one’s quality of life. Also, analyzing a combination of FIM measures alongside other outcome measures collected by the TBIMS database, such as the Glasgow Outcome Scale–Extended, the Supervision Rating Scale, and the PART-O participation instrument, would yield a more robust, multidimensional assessment of long-term recovery trajectories and societal reintegration.

Lastly, with the association of TBIs from violence and assaults having poorer outcomes, to understand these disparities more deeply, it is imperative to incorporate psychosocial factors into future models and analyses. The TBIMS database collects information regarding substance use history, arrests, and pre- and post-injury psychiatric issues. Including these variables in future models as possible confounding or mediating variables in analyses of assault/violence-related TBI outcomes may help to elucidate the complex relationships between the mechanism of TBI, psychological variables, and functional recovery, offering insights into optimizing targeted rehabilitation protocols.

## CONCLUSION

The mechanism of TBI significantly impacts long-term functional outcomes during recovery, with violence-related mechanisms (particularly gunshot wounds) leading to poorer locomotor independence over an extended time. The significant negative estimates shown in Figure 1 suggest that TBIs caused by motor vehicles, gunshots, assaults, other violence, and pedestrian activities may be disproportionately more likely to experience difficulties with their long-term locomotion.

The study findings are clinically significant as they highlight groups who may have a disproportionately higher risk for experiencing impairments, enabling clinicians to design and develop targeted rehabilitation plans and interventions to counteract these disabilities, thereby reducing the significant burden facing individuals, families, and society following a TBI. The combined use of Rasch analysis and linear mixed-effects modeling significantly enhanced the analytical rigor of this study. The analysis transforms raw FIM scores into interval-level logit scores, addressing potential ceiling effects and yielding a more accurate assessment of the latent functional ability. Coupling the Rasch analysis with linear mixed-effects regression effectively accommodates the complex, heterogeneous longitudinal TBI data and captures the fixed effect of TBI mechanisms as well as individual-level variability in recovery trajectories, revealing nuanced differences that may have gone unnoticed in previous research.

To advance understanding and optimize recovery outcomes, future work should investigate the impact of psychosocial characteristics, examine nonlinear recovery patterns using modeling techniques such as spline models, and explore analyses of long-term cognitive functional outcomes. This will offer a more complete and accurate depiction of long-term TBI recovery trajectories.

## Data Availability

No Available Data.

*Appendices*

## References

1. James SL, Theadom A, Ellenbogen RG, Bannick MS, Montjoy-Venning W, Lucchesi LR, et al. Global, regional, and national burden of traumatic brain injury and spinal cord injury, 1990–2016: a systematic analysis for the Global Burden of Disease Study 2016. The Lancet Neurology. 2019;18(1):56–87. 10.1016/S1474-4422(18)30415-0

2. Yue J, Deng H. Traumatic Brain Injury: Contemporary Challenges and the Path to Progress. JCM. 2023 May 5;12(9):3283. doi:10.3390/jcm12093283

3. Zhong H, Feng Y, Shen J, Rao T, Dai H, Zhong W, et al. Global Burden of Traumatic Brain Injury in 204 Countries and Territories From 1990 to 2021. American Journal of Preventive Medicine. 2025 Apr;68(4):754–63. doi:10.1016/j.amepre.2025.01.001

4. Demir Y. Factors affecting functional outcome in patients with traumatic brain injury sequelae: Our single-center experiences on brain injury rehabilitation. Turk J Phys Med Rehab. 2019 Mar 8;65(1):67–73. doi:10.5606/tftrd.2019.2281

5. Maas AIR, Menon DK, Manley GT, Abrams M, Åkerlund C, Andelic N, et al. Traumatic brain injury: progress and challenges in prevention, clinical care, and research. The Lancet Neurology. 2022 Nov;21(11):1004–60. doi:10.1016/S1474-4422(22)00309-X

6. Van Dijck JTJM, Dijkman MD, Ophuis RH, De Ruiter GCW, Peul WC, Polinder S. In-hospital costs after severe traumatic brain injury: A systematic review and quality assessment. Kerezoudis P, editor. PLoS ONE. 2019 May 9;14(5):e0216743. doi:10.1371/journal.pone.0216743

7. Rasmussen MS, Zhang Y, Andelic N, Aas E. Health care costs and service utilization in the first year following moderate to severe traumatic injury. BMC Health Serv Res. 2024 Dec 3;24(1):1535. doi:10.1186/s12913-024-12016-6

8. Vardon-Bounes F, Gracia R, Abaziou T, Crognier L, Seguin T, Labaste F, et al. A study of patients’ quality of life more than 5 years after trauma: a prospective follow-up. Health Qual Life Outcomes. 2021 Dec;19(1):18. doi:10.1186/s12955-020-01652-1

9. Andelic N, Arango-Lasprilla JC, Perrin PB, Sigurdardottir S, Lu J, Landa LO, et al. Modeling of Community Integration Trajectories in the First Five Years after Traumatic Brain Injury. Journal of Neurotrauma. 2016 Jan;33(1):95–100. doi:10.1089/neu.2014.3844

10. Maas AIR, Menon DK, Adelson PD, Andelic N, Bell MJ, Belli A, et al. Traumatic brain injury: integrated approaches to improve prevention, clinical care, and research. The Lancet Neurology. 2017 Dec;16(12):987–1048. doi:10.1016/S1474-4422(17)30371-X

11. Butcher I, McHugh GS, Lu J, Steyerberg EW, Hernández AV, Mushkudiani N, et al. Prognostic Value of Cause of Injury in Traumatic Brain Injury: Results from The IMPACT Study. Journal of Neurotrauma. 2007 Feb;24(2):281–6. doi:10.1089/neu.2006.0030

12. Majdan M, Mauritz W, Brazinova A, Rusnak M, Leitgeb J, Janciak I, et al. Severity and outcome of traumatic brain injuries (TBI) with different causes of injury. Brain Injury. 2011 Aug;25(9):797–805. doi:10.3109/02699052.2011.581642

13. Howrey BT, Graham JE, Pappadis MR, Granger CV, Ottenbacher KJ. Trajectories of Functional Change After Inpatient Rehabilitation for Traumatic Brain Injury. Archives of Physical Medicine and wRehabilitation. 2017 Aug;98(8):1606–13. doi:10.1016/j.apmr.2017.03.009

14. Colantonio A, Ratcliff G, Chase S, Kelsey S, Escobar M, Vernich L. Long term outcomes after moderate to severe traumatic brain injury. Disability and Rehabilitation. 2004 Mar 4;26(5):253–61. doi:10.1080/09638280310001639722

15. Ruet A, Bayen E, Jourdan C, Ghout I, Meaude L, Lalanne A, et al. A Detailed Overview of Long-Term Outcomes in Severe Traumatic Brain Injury Eight Years Post-injury. Front Neurol. 2019 Feb 21;10:120. doi:10.3389/fneur.2019.00120

16. Pasipanodya EC, Teranishi R, Dirlikov B, Duong T, Huie H. Characterizing Profiles of TBI Severity: Predictors of Functional Outcomes and Well-Being. Journal of Head Trauma Rehabilitation. 2023 Jan;38(1):E65–78. doi:10.1097/HTR.0000000000000791

17. Cappa KA, Conger JC, Conger AJ. Injury severity and outcome: A meta-analysis of prospective studies on TBI outcome. Health Psychology. 2011;30(5):542–60. doi:10.1037/a0025220

18. Lafta G, Sbahi H. Factors associated with the severity of traumatic brain injury. Medicine and Pharmacy Reports. 2023 Jan 18;96(1):58–64. doi:10.15386/mpr-2314

19. Corrigan F, Wee IC, Collins-Praino LE. Chronic motor performance following different traumatic brain injury severity—A systematic review. Front Neurol. 2023 May 11;14:1180353. doi:10.3389/fneur.2023.1180353

20. Bertozzi G, Maglietta F, Sessa F, Scoto E, Cipolloni L, Di Mizio G, et al. Traumatic brain injury: a forensic approach: a literature review. Current neuropharmacology. 2020;18(6):538–50.

21. Dijkers MP, Harrison-Felix C, Marwitz JH. The Traumatic Brain Injury Model Systems: History and Contributions to Clinical Service and Research. JOURNAL OF HEAD TRAUMA REHABILITATION. 2010.

22. Tso S, Saha A, Cusimano MD. The Traumatic Brain Injury Model Systems National Database: A Review of Published Research. Neurotrauma Reports. 2021 Jan 1;2(1):149–64. doi:10.1089/neur.2020.0047

23. Traumatic Brain Injury Model Systems National Data and Statistical Center. TBI Model Systems National Database: 2021 profile of people within the Traumatic Brain Injury Model Systems [Internet]. Englewood, Colorado, USA: Craig Hospital; 2025 [cited 2025 Aug 26]. Available from: https://www.tbindsc.org

24. Hall K, Cope DN, Rappaport M. Glasgow Outcome Scale and Disability Rating Scale: comparative usefulness in following recovery in traumatic head injury. Archives of physical medicine and rehabilitation. 1985;66(1):35–7.

25. Rappaport M, Hall K, Hopkins K, Belleza T, Cope DN. Disability rating scale for severe head trauma: coma to community. Archives of physical medicine and rehabilitation. 1982;63(3):118–23.

26. Jennett B, Snoek J, Bond MR, Brooks N. Disability after severe head injury: observations on the use of the Glasgow Outcome Scale. Journal of Neurology, Neurosurgery & Psychiatry. 1981 Apr 1;44(4):285–93. doi:10.1136/jnnp.44.4.285

27. Wilson JTL, Pettigrew LEL, TTeasdale GM. Structured Interviews for the Glasgow Outcome Scale and the Extended Glasgow Outcome Scale: Guidelines for Their Use. Journal of Neurotrauma. 1998 Aug 1;15(8):573–85. doi:10.1089/neu.1998.15.573

28. Boake C. Supervision Rating Scale: a measure of functional outcome from brain injury. Archives of physical medicine and rehabilitation. 1996;77(8):765–72.

29. Dijkers M, Cicerone K, Heinemann A, Brown M, Whiteneck G. Poster 89: PART-O: A New Measure of Satisfaction with Participation. Archives of Physical Medicine and Rehabilitation. 2009 Oct 1;90(10):e39. doi:10.1016/j.apmr.2009.08.123

30. Diener E, Emmons RA, Larsen RJ, Griffin S. The satisfaction with life scale. Journal of personality assessment. 1985;49(1):71–5.

31. Dams-O’Connor K, Juengst SB, Bogner J, Chiaravalloti ND, Corrigan JD, Giacino JT, et al. Traumatic brain injury as a chronic disease: insights from the United States Traumatic Brain Injury Model Systems Research Program. The Lancet Neurology. 2023 Jun;22(6):517–28. doi:10.1016/S1474-4422(23)00065-0

32. Granger CV, Hamilton BB, Linacre JM, Heinemann AW, Wright BD. Performance profiles of the functional independence measure. American journal of physical medicine & rehabilitation. 1993;72(2):84–9.

33. Keith R. The functional independence measure; a new tool for rehabilitation. Adv Clin Rehabil. 1987;1:6–18.

34. Pretz CR, Kean J, Heinemann AW, Kozlowski AJ, Bode RK, Gebhardt E. A Multidimensional Rasch Analysis of the Functional Independence Measure Based on the National Institute on Disability, Independent Living, and Rehabilitation Research Traumatic Brain Injury Model Systems National Database. Journal of Neurotrauma. 2016 Jul 15;33(14):1358–62. doi:10.1089/neu.2015.4138

35. Donaghy S, Wass PJ. Interrater reliability of the functional assessment measure in a brain injury rehabilitation program. Archives of Physical Medicine and Rehabilitation. 1998 Oct;79(10):1231–6. doi:10.1016/S0003-9993(98)90267-2

36. Fisher RA. Statistical methods for research workers. 1934.

37. R Core Team. R [Internet]. 2025. (R: A language and environment for statistical computing). Available from: https://www.R-project.org/

38. Robitzsch A, Kiefer T, Wu M. TAM: Test Analysis Modules [Internet]. 2025 [cited 2025 Nov 13]. Available from: https://cran.r-project.org/web/packages/TAM/index.html

39. Pruim R, Kaplan DT, Horton NJ. The mosaic Package: Helping Students to “Think with Data” Using R. The R Journal. 2017;9(1):77–102.

40. Bates D, Mächler M, Bolker B, Walker S. Fitting Linear Mixed-Effects Models Using lme4. 2015.

41. Wickham H. ggplot2: Elegant Graphics for Data Analysis [Internet]. Springer-Verlag New York; 2016. Available from: https://ggplot2.tidyverse.org

42. Alao T, Munakomi S, Waseem M. Penetrating Head Trauma. In. Treasure Island (FL): StatPearls Publishing; 2025 [cited 2026 Jan 17]. Available from: http://www.ncbi.nlm.nih.gov/books/NBK459254/ PubMed PMID: 29083824.

43. Cordingley DM, Marquez I, Buchwald SC, Zeiler FA. Does Physical Fitness Prior to Traumatic Brain Injury Affect Recovery Outcomes? A Scoping Review of Human and Animal Research. Neurotrauma Reports. 2025;6(1):768.

44. Taylor JM, Montgomery MH, Gregory EJ, Berman NEJ. Exercise preconditioning improves traumatic brain injury outcomes. Brain Research. 2015 Oct;1622:414–29. doi:10.1016/j.brainres.2015.07.009

45. Rouquette A, Hardouin JB, Vanhaesebrouck A, Sébille V, Coste J. Differential Item Functioning (DIF) in composite health measurement scale: Recommendations for characterizing DIF with meaningful consequences within the Rasch model framework. PloS one. 2019;14(4):e0215073.

46. Nudo RJ. Recovery after brain injury: mechanisms and principles. Front Hum Neurosci. 2013;7. doi:10.3389/fnhum.2013.00887

47. Zotey V, Andhale A, Shegekar T, Juganavar A. Adaptive Neuroplasticity in Brain Injury Recovery: Strategies and Insights. Cureus. 2023 Sep 24. doi:10.7759/cureus.45873

